# Lower Social Support is Associated with Accelerated Epigenetic Aging: Results from the Health and Retirement Study

**DOI:** 10.1101/2022.06.03.22275977

**Authors:** Kelly E. Rentscher, Eric T. Klopack, Eileen M. Crimmins, Teresa E. Seeman, Steve W. Cole, Judith E. Carroll

**Affiliations:** Department of Psychiatry and Behavioral Medicine, Medical College of Wisconsin, Milwaukee, WI, 53226, USA; Cousins Center for Psychoneuroimmunology, Semel Institute for Neuroscience and Human Behavior, University of California, Los Angeles, Los Angeles, CA, 90095, USA; Leonard Davis School of Gerontology, University of Southern California, Los Angeles, CA, 90089, USA; Division of Geriatrics, David Geffen School of Medicine, University of California, Los Angeles, CA, 90095, USA

**Author notes:** **Corresponding author:** Kelly E. Rentscher, Ph.D., Department of Psychiatry and Behavioral Medicine, Medical College of Wisconsin, 1000 N. 92^nd^ St. Milwaukee, WI 53226. **Email addresses for other authors:** Eric T. Klopack Eileen M. Crimmins Teresa E. Seeman Steve W. Cole Judith E. Carroll.

**Keywords:** social support, social strain, biological aging, epigenetic age, DNA methylation

## Abstract

Growing evidence suggests that social relationship quality can influence age-related health outcomes, although how the quality of one’s relationships directly relates to the underlying aging process is less clear. We hypothesized that lower social support and higher relationship strain would be associated with an accelerated epigenetic aging profile among older adults in the Health and Retirement Study. Adults (*N* = 3,647) aged 50-96 years completed ratings of support and strain in relationships with their spouse, children, other family, and friends. They also provided a blood sample and DNA methylation profiling derived epigenetic aging measures: Horvath, Hannum, PhenoAge, GrimAge, and Dunedin Pace of Aging methylation (PoAm). Generalized linear models adjusting for age, sex, and race/ethnicity revealed that lower support from one’s spouse, children, other family, and friends and higher strain with one’s spouse, children, and friends was associated with an accelerated epigenetic aging profile. In secondary analyses that further adjusted for socioeconomic and lifestyle factors and following false discovery rate correction, support from other family members and friends was associated with epigenetic aging. Findings suggest that lower support within close relationships relates to epigenetic aging acceleration, offering one mechanism through which relationship quality might influence risk for age-related disease.

## 1. Introduction

A sizeable literature has established that the quality of one’s social relationships can have a significant impact on health and well-being across the lifespan [1,2]. Current theoretical frameworks posit that social relationships promote health by fulfilling basic needs for social connection and by providing a buffering resource during times of stress; however, they can also be a source of conflict and strain [3–7]. Research has identified perceived social support and social strain as distinct dimensions of social relationship quality that can influence health [3,6– 9]. Whereas social support is defined as the availability of resources, advice, understanding, or acceptance [3,10], social strain has been described as criticism, insensitivity, demands, or feeling let down within one’s current relationships [6,11]. Social support and strain are concurrently and prospectively associated with age-related declines and disease in middle-aged and older adults, including functional limitations [12,13], poorer physical and cognitive functioning [14–16], greater number of chronic conditions [12], and incidence and progression of cardiovascular disease [17–20]. They are also reliable predictors of all-cause mortality [1,21–24] and mortality from cancer [25,26], stroke [27], and cardiovascular disease [23,28]. Together, this research suggests that social support and strain influence aging, although how these processes directly relate to the underlying aging process is less clear.

Epigenetic aging offers one approach to track the underlying aging process. Epigenetic aging refers to age-related alterations to the epigenome (i.e., chemical compounds that modify DNA but do not change its coding sequence), which include histone modifications, chromatin remodeling, and changes in DNA methylation patterns [29]. One of the most widely used approaches to measuring epigenetic aging, termed the ‘epigenetic clock,’ was developed by identifying distinct regions of the DNA that become hypo- or hyper-methylated with age, and correlates with chronological age across a range of cell types and tissues [30–32]. More recent versions of the epigenetic clock were developed based on DNA methylation patterns that are associated with multiple aging biomarkers and are predictive of phenotypic aging outcomes (PhenoAge) [33] and time to death (GrimAge) [34]. In contrast to the epigenetic clock measures, the Dunedin Pace of Aging methylation (PoAm) measure was developed to estimate an individual’s *rate* of biological aging at a single point in time, based on data from the Dunedin cohort and changes in 18 biomarkers of organ-system integrity assessed over a 12-year period [35]. Accelerated epigenetic aging, as assessed by these measures, has been associated with multiple age-related conditions, including frailty [36–38], physical limitations [34,35,39–42], cognitive decline [35,41,43,44], cancer [34,42,45–50], diabetes [34,50], stroke [51,52], and cardiovascular disease [34,42,50], as well as all-cause [34,35,58–60,37,42,50,53–57] and specific-cause (e.g., cancer; [42,47,49,57]) mortality.

The present study aimed to investigate whether perceived social support and strain were associated with accelerated epigenetic aging in a nationally representative sample older adults in the Health and Retirement Study (HRS). As part of the 2016 self-administered psychosocial questionnaire, participants completed ratings of support and strain in relationships with their spouse, children, other family members, and friends. As part of the 2016 Venous Blood Study, participants also provided a blood sample and DNA methylation profiling was performed to derive epigenetic aging measures. This investigation focused on five established, *a priori*-identified epigenetic measures: the Horvath, Hannum, PhenoAge, and GrimAge clocks, and the Dunedin PoAm. Based on previous research linking social relationship quality to age-related conditions, we hypothesized that higher perceived support within close relationships would be associated with a younger epigenetic aging profile, characterized by a younger Horvath, Hannum, PhenoAge, and GrimAge epigenetic age and a slower Dunedin PoAm. We also expected that greater strain within close relationships would be associated with an accelerated epigenetic aging profile, characterized by an older Horvath, Hannum, PhenoAge, and GrimAge epigenetic age and a faster Dunedin PoAm in older adulthood.

## 2. Results

### 2.1. Preliminary analyses

Table 1 presents descriptive statistics for the social and epigenetic aging variables. On average, participants reported more support than strain for all relationship types. The average epigenetic age for the clock measures (Horvath, Hannum, PhenoAge, GrimAge) ranged from 54.04 to 67.20. The average Dunedin PoAm was 1.07 years of epigenetic age for each year of chronological age.

**Table 1.**
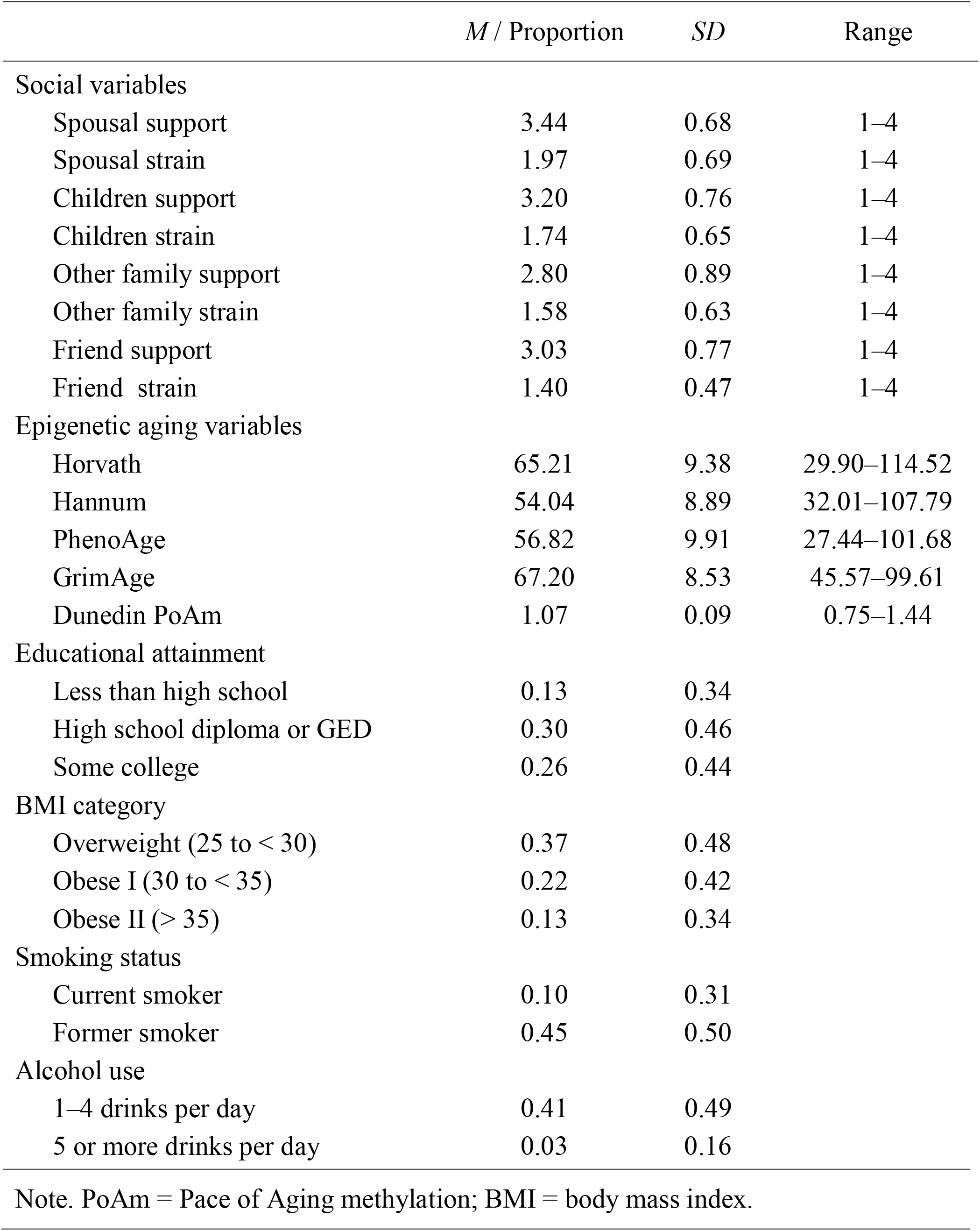
Descriptive statistics for the main study variables.

|  | *M* / Proportion | *SD* | Range |
| --- | --- | --- | --- |
| Social variables |  |  |  |
| Spousal support | 3.44 | 0.68 | 1–4 |
| Spousal strain | 1.97 | 0.69 | 1–4 |
| Children support | 3.20 | 0.76 | 1–4 |
| Children strain | 1.74 | 0.65 | 1–4 |
| Other family support | 2.80 | 0.89 | 1–4 |
| Other family strain | 1.58 | 0.63 | 1–4 |
| Friend support | 3.03 | 0.77 | 1–4 |
| Friend strain | 1.40 | 0.47 | 1–4 |
| Epigenetic aging variables |  |  |  |
| Horvath | 65.21 | 9.38 | 29.90–114.52 |
| Hannum | 54.04 | 8.89 | 32.01–107.79 |
| PhenoAge | 56.82 | 9.91 | 27.44–101.68 |
| GrimAge | 67.20 | 8.53 | 45.57–99.61 |
| Dunedin PoAm | 1.07 | 0.09 | 0.75–1.44 |
| Educational attainment |  |  |  |
| Less than high school | 0.13 | 0.34 |  |
| High school diploma or GED | 0.30 | 0.46 |  |
| Some college | 0.26 | 0.44 |  |
| BMI category |  |  |  |
| Overweight (25 to < 30) | 0.37 | 0.48 |  |
| Obese I (30 to < 35) | 0.22 | 0.42 |  |
| Obese II (> 35) | 0.13 | 0.34 |  |
| Smoking status |  |  |  |
| Current smoker | 0.10 | 0.31 |  |
| Former smoker | 0.45 | 0.50 |  |
| Alcohol use |  |  |  |
| 1–4 drinks per day | 0.41 | 0.49 |  |
| 5 or more drinks per day | 0.03 | 0.16 |  |

Note. PoAm = Pace of Aging methylation; BMI = body mass index.

### 2.2. Support and strain with spouse

Generalized linear models examined spousal support and strain as separate predictors of epigenetic aging variables, adjusting for age, sex, and race/ethnicity. Consistent with hypotheses, higher perceived spousal support was associated with a younger Hannum epigenetic age, PhenoAge, and GrimAge, and a slower Dunedin PoAm (Figure 1; Supplemental Information Table 1). A one-point increase on the spousal support scale was associated with a Hannum age 0.38 years younger, a PhenoAge 0.61 years younger, a GrimAge 0.34 years younger, and a Dunedin pace of aging 0.008 years younger per chronological year than same-aged peers. Spousal support was not associated with the Horvath clock. In secondary analyses that adjusted for educational attainment and lifestyle factors (BMI category, smoking status, and alcohol use), associations between spousal support and the Hannum, GrimAge and Dunedin PoAm measures were reduced to non-significance; only the association between spousal support and PhenoAge remained statistically significant (Supplemental Information Table 1). However, this association was not significant following 5% false discovery rate (FDR) correction for multiple testing. Also as expected, greater spousal strain was associated with an accelerated Hannum epigenetic age (Figure 2; Supplemental Information Table 2). A one-point increase in spousal strain was associated with a Hannum age 0.35 years older than same aged peers. In secondary analyses, this association remained statistically significant after adjusting for educational attainment and lifestyle factors but was not significant following FDR correction for multiple testing (Supplemental Information Table 2). Spousal strain was not associated with the Horvath, PhenoAge, GrimAge, or Dunedin PoAm measures.

**Legend for Figure 1.**
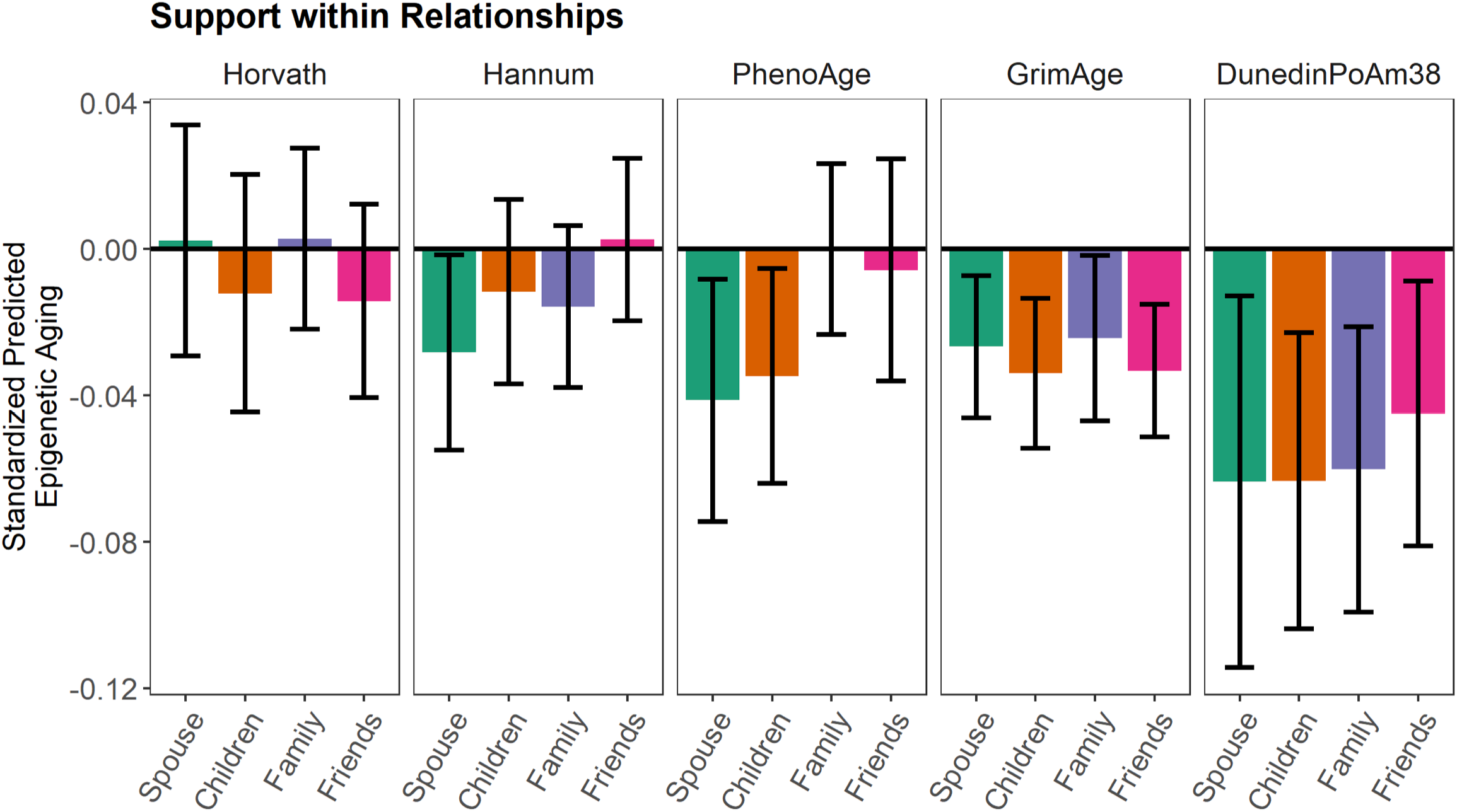
Standardized coefficients for the association between social support within spousal, child, other family, and friend relationships and epigenetic aging measures, adjusting for age, sex, and race/ethnicity. Standard error bars represent 95% confidence intervals for each point estimate.

**Legend for Figure 2.**
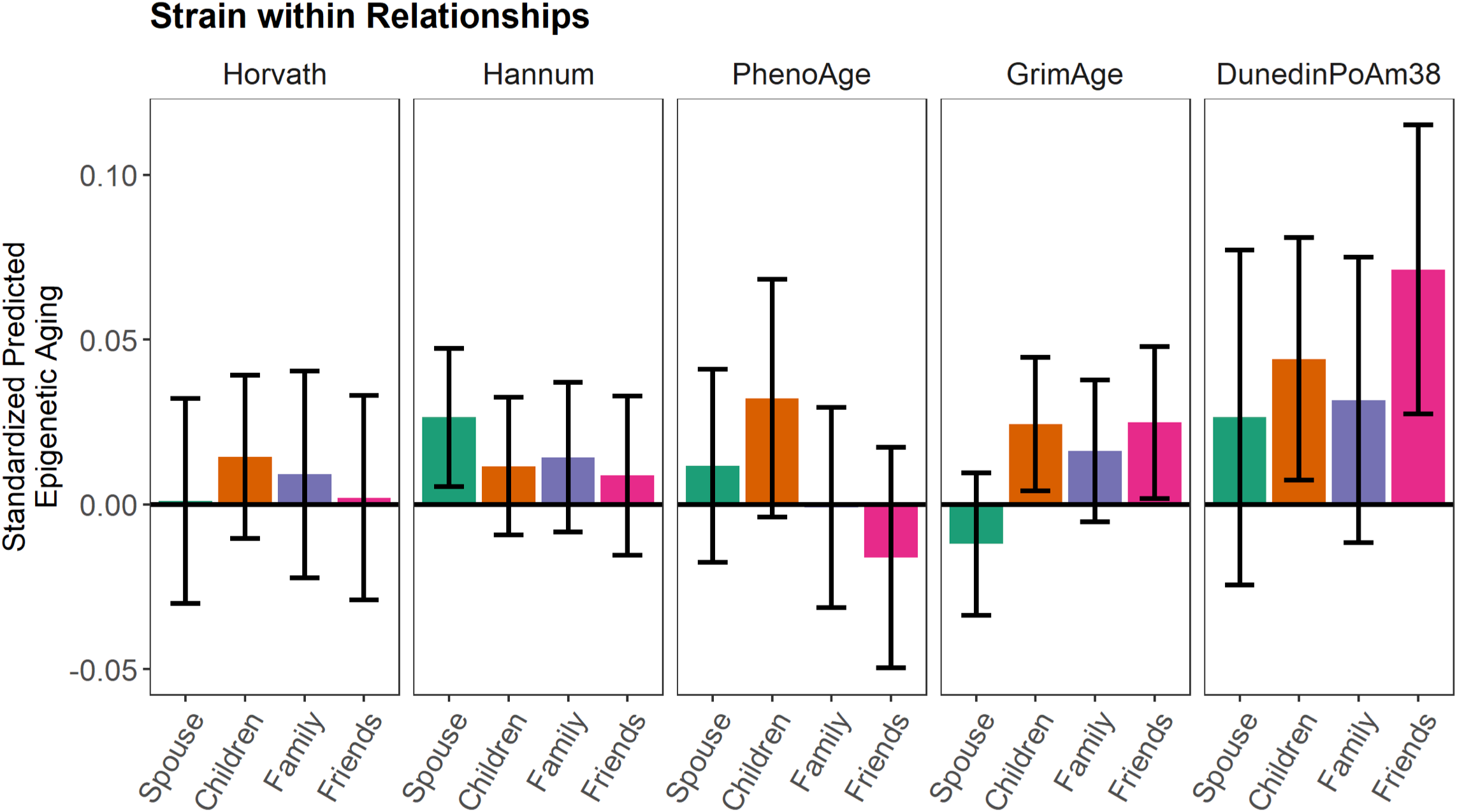
Standardized coefficients for the association between social strain within spousal, child, other family, and friend relationships and epigenetic aging measures, adjusting for age, sex, and race/ethnicity. Standard error bars represent the 95% confidence intervals for each point estimate.

### 2.3. Support and strain with children

Consistent with hypotheses, higher support from one’s children was associated with a younger PhenoAge and GrimAge, and a slower Dunedin PoAm (Figure 1; Supplemental Information Table 3). A one-point increase in support from one’s children was associated with a PhenoAge 0.47 years younger, a GrimAge 0.39 years younger, and a Dunedin pace of aging 0.008 years younger per chronological year than same-aged peers. In secondary analyses, the association between support from one’s children and PhenoAge was reduced to non-significance after adjusting for educational attainment and lifestyle factors, but associations with GrimAge and Dunedin PoAm remained statistically significant (Supplemental Information Table 3). However, these associations were reduced to non-significance following FDR correction for multiple testing. Support from one’s children was not associated with the Horvath or Hannum clocks. Also as expected, greater strain with one’s children was associated with an accelerated GrimAge and faster Dunedin PoAm (Figure 2; Supplemental Information Table 4). A one-point increase in strain with one’s children was associated with a GrimAge 0.32 years older, and a Dunedin pace of aging 0.006 years older per chronological year than same-aged peers. However, in secondary analyses, these associations were reduced to non-significance after adjusting for educational attainment and lifestyle factors and FDR correction (Supplemental Information Table 4). Strain with children was not associated with the Horvath, Hannum, or PhenoAge measures.

### 2.4. Support and strain with other family members

Consistent with hypotheses, higher support from other family members was associated with a younger GrimAge and slower Dunedin PoAm (Figure 1; Supplemental Information Table 5). A one-point increase in support from other family members was associated with a GrimAge 0.24 years younger, and a Dunedin pace of aging 0.006 years younger per chronological year than same-aged peers. In secondary analyses, these associations remained statistically significant after adjusting for educational attainment and lifestyle factors (Supplemental Information Table 5). The association between support from other family members and Dunedin PoAm also remained significant following FDR correction. Support from other family members was not associated with the Horavth, Hannum, or PhenoAge measures. Strain with other family members was not associated with any of the epigenetic aging measures (Figure 2; Supplemental Information Table 6).

### 2.5. Support and strain with friends

Consistent with hypotheses, higher support from friends was associated with a younger GrimAge and slower Dunedin PoAm (Figure 1; Supplemental Information Table 7). A one-point increase in support from friends was associated with a GrimAge 0.38 years younger, and a Dunedin pace of aging 0.005 years younger per chronological year than same-aged peers. In secondary analyses, the association between support from friends and Dunedin PoAm was reduced to non-significant after adjusting for educational attainment and lifestyle factors, but the association with GrimAge remained statistically significant (Supplemental Information Table 7). The association between support from friends and GrimAge also remained statistically significant following FDR correction. Support from friends was not associated with the Horvath, Hannum, or PhenoAge measures. Also as expected, greater strain with friends was associated with an accelerated GrimAge and faster Dunedin PoAm (Figure 2; Supplemental Information Table 8). A one-point increase in strain with friends was associated with a GrimAge 0.43 years older and a Dunedin pace of aging 0.013 years older per chronological year than same-aged peers. In secondary analyses, the associations between strain with friends and GrimAge and Dunedin PoAm were no longer statistically significant after adjusting for educational attainment and lifestyle factors and FDR correction (Supplemental Information Table 8). Strain within friendships was not associated with the Horvath, Hannum, or PhenoAge measures.

## 3. Discussion

The present study investigated associations between social relationship quality and epigenetic aging in a nationally representative sample of older adults in the Health and Retirement Study. Specifically, we examined whether lower perceived support and higher strain in relationships with one’s spouse, children, other family members, and friends was concurrently related to an accelerated epigenetic aging profile. As hypothesized, individuals who reported feeling understood, that they could rely upon, or that they could open up to their spouse, children, family members, and friends had a younger epigenetic aging profile relative to same-aged peers. Also as expected, individuals who reported that their spouse, children, and friends made too many demands on them, criticized them, let them down, or got on their nerves had an accelerated aging profile relative to same-aged peers. On average, the difference in epigenetic aging between individuals who reported the lowest and highest levels of social support ranged from 0.72 to 1.83 years, depending on the relationship type and the epigenetic measure. The average difference in epigenetic aging between individuals who reported the lowest and highest levels of social strain ranged from 0.42 to 1.29 years, depending on the relationship type and the epigenetic measure. It was somewhat surprising that social support and strain were less consistently or not at all associated with the Horvath and Hannum clocks, although it is noteworthy that both of these measures were developed to predict chronological age and may be capturing different aspects of the aging process than the PhenoAge, GrimAge, and Dunedin PoAm measures [50].

In secondary analyses that further adjusted for educational attainment and lifestyle factors that previous research found were associated with epigenetic aging [34,42,50,61,62], social support was more robustly associated with epigenetic aging than social strain. Specifically, there were unique associations between spousal support and PhenoAge, support from children and other family members and GrimAge and Dunedin PoAm, and support from friends and GrimAge that were over and above these well-established health risk factors. On the other hand, only spousal strain was uniquely associated with the Hannum clock. However, it is important to note that, following false discovery rate correction for multiple testing, only support from other family members and friends were associated with Dunedin PoAm and GrimAge, respectively. These findings are particularly noteworthy in this sample of older adults who may be experiencing declines in their health, as PhenoAge, GrimAge, and Dunedin PoAm are predictive of multiple age-related conditions such as cognitive decline, loss of physical function, cancer, cardiovascular disease, and time to death [34,42]. In addition, researchers have found that relationships with close others become more salient with age [63,64], and in some cases, they may become more stable or involuntary (e.g., more difficult to choose to exit) as older adults who experience health declines may rely more on others for support [65]. Therefore, perceiving that close relationships with family and friends are lower in support and/or higher in conflict or strain may have a particular influence on health and well-being with increasing age—and these findings suggest that epigenetic aging may be one mechanism through which this occurs.

These findings are consistent with and contribute to a growing literature on the influence of social relationship quality on key biological aging processes. For instance, several studies have linked lower levels of social support and higher strain to increased activation of transcription factor NF-κB, which regulates the expression of inflammatory genes [66], and peripheral markers of inflammation (e.g., IL-6, IL-8, CRP, TNF-alpha) in middle-aged and older adults [67–74]. Other research has suggested that lower social support and a greater number of ambivalent relationships (i.e., characterized by high positivity and negativity) are associated with shorter telomere length [75–77]. The present findings also extend an emerging literature on psychosocial stress and epigenetic aging, with several studies linking early life adversity [78– 84], lower socioeconomic status [79,81,85–89], and traumatic stress [90,91] to an accelerated epigenetic aging profile. As low perceived social support and social strain have been considered a form of social stress, social relationship quality may affect epigenetic aging through similar mechanisms; however, the specific cellular and molecular mechanisms through which this might occur remain largely unknown. Chronic or repeated activation of the sympathetic nervous system (SNS) and the hypothalamus-adrenal-pituitary (HPA) axis in response to stress releases neuroendocrine mediators (e.g., catecholamines, glucocorticoids) that interact with receptors on the surface of cells [92]. Mounting evidence suggests that this stress signaling cascade can initiate various physiological processes within cells, including those that contribute to accelerated biological aging, such as cell stress, DNA damage, inflammation, and mitochondrial dysfunction (Polsky, Rentscher, & Caroll, under revision). It will be important for future research to begin to delineate the biological pathways through which exposure to social adversity and associated neuroendocrine mediators may modify these DNA methylation patterns and other epigenetic processes to alter rates of aging.

This study has several limitations, which suggest directions for future research. Most notably, the single-occasion measurement of epigenetic aging limited our analysis to concurrent associations and precluded the investigation of the influence of social relationship quality on changes in epigenetic aging over time. On account of this, we were also unable to test the alternative hypothesis that changes in epigenetic aging, as a marker of an underlying aging process, might influence shifts in social relationship quality. Future research would benefit from testing the directionally of the observed effects and linking these associations to age-related health outcomes in later life. In addition, the social support measures for this study focused primarily on emotional support and did not address other forms of support, such as tangible or informational support.

Despite these limitations, this is the first study to demonstrate that lower perceived support in relationships with family members and friends are uniquely associated with an accelerated epigenetic aging profile in older adulthood, over and above well-established lifestyle factors such as smoking status and alcohol use. These findings extend previous research by identifying epigenetic aging as a biological aging mechanism through which social relationship quality might influence aging and age-related health outcomes such as cancer, cardiovascular disease, dementia, and early mortality. In addition, the present sample was relatively large, socioeconomically diverse, and nationally representative of the U.S. population of older adults, which increases generalizability of the findings. Importantly, in light of emerging evidence that these epigenetic aging mechanisms may be sensitive to and modifiable by behavioral intervention [93,94], these findings suggest that close relationship quality—particularly with family members and friends—may represent a behavioral target for intervention in older adulthood that has the potential to prevent, slow, or reverse accelerated aging and extend the healthspan (number of years free from age-related disease and disability) and lifespan.

## 4. Methods

### 4.1. Ethics Statement

This investigation has been conducted in accordance with the ethical standards and according to the Declaration of Helsinki and according to national and international guidelines and has been approved by the Institutional Review Board at the University of Michigan.

### 4.2. Participants

The present study used data from the University of Michigan Health and Retirement Study (HRS), a longitudinal, nationally representative study of nearly 20,000 U.S. adults over the age of 50 [95]. For this study, participants were 4,018 adults aged 50-96 years who provided a blood sample as part of the 2016 Venous Blood Study (VBS) that was used to assess epigenetic aging [96]. The epigenetic aging subsample of the VBS was designed to be representative of the U.S. population when weighted. For the present analysis, 285 participants were missing data for at least one social support or strain variable and an additional 86 participants were missing data for at least one covariate; therefore, the final sample size was 3,647. The weighted sample had a median age of 68.7 years and was 55.1% female. Participants were non-Hispanic white (79.5%), non-Hispanic Black (9.2%), Hispanic (7.9%), and non-Hispanic of another race (3.3%) respondents. The educational distribution of the participants included less than a high school education (13.1%), high school diploma or GED (30.2%), some college (26.0%), and college diploma or higher (30.7%).

### 4.3. Procedures

Participants in the HRS study complete core interviews every other year, and self-administered psychosocial questionnaires are also given to alternating random halves of the full sample every two years. As part of the 2016 self-administered psychosocial questionnaire, participants completed ratings of support and strain in their relationships with their spouse, children, other family members, and friends. If participants were missing social support and strain data from 2016, we used data from the 2014 questionnaire. If they were missing data from 2016 and 2014, we used data from the 2012 questionnaire, and so forth, to 2008 (when the social support and strain measures were first included in the questionnaire). Participants also provided a blood sample as part of the 2016 Venous Blood Study (VBS) and DNA methylation profiling was performed using the Illumina Infinium Methylation EPIC BeadChip (Illumina, San Diego, CA) to derive the epigenetic aging measures, as described in detail previously [96,97].

### 4.4. Measures

#### 4.4.1. Social support and strain measures

Participants completed ratings of perceived support and strain within four types of relationships: their spouse, children, other family, and friends. For each relationship type, three items assessed support: “How much do they really understand the way you feel about things?”, “How much can you rely on them if you have a serious problem?”, and “How much can you open up to them if you need to talk about your worries?”. Four items assessed strain: “How often do they make too many demands on you?”, “How much do they criticize you?”, “How much do they let you down when you are counting on them?”, and “How much do they get on your nerves?”. Responses ranged from 1 (*not at all*) to 4 (*a lot*). Items were averaged to create a composite score, with higher scores indicating greater support or strain.

#### 4.4.2. Epigenetic aging measures

The epigenetic aging measures for this study included the Horvath, Hannum, PhenoAge, and GrimAge clocks and the Dunedin Pace of Aging methylation (PoAm). The Horvath estimate of epigenetic age is based on DNA methylation levels at 353 cytosine-phosphate-guanine base pair (CpG) sites and was developed as a predictor of chronological age across multiple tissues and cell types [31]. The Hannum estimate of epigenetic age is based on DNA methylation levels at 71 CpG sites and was developed as a predictor of chronological age in whole blood samples [32]. Phenotypic epigenetic age—also referred to as PhenoAge—is estimated from DNA methylation levels at 513 CpG sites and was developed as a predictor of mortality risk based on 9 markers of tissue and immune function (albumin, creatinine, serum glucose, C-reactive protein [CRP], lymphocyte percent, mean (red) cell volume, red cell distribution width, alkaline phosphatase, and white blood cell count) and chronological age in whole blood samples [42]. GrimAge is estimated from DNA methylation levels at 1,030 total CpG sites and was developed as a predictor of time to death based on 7 DNA methylation surrogates of plasma proteins associated with physiological risk and stress factors (adrenomedullin, beta-2 microglobulin, cystatin C, growth differentiation factor 15 [GDF-15], leptin, plasminogen activation inhibitor 1 [PAI-1], tissue inhibitor metalloproteinase 1 [TIMP-1]) and a DNA methylation-based estimator of smoking pack years [34]. Dunedin Pace of Aging methylation (PoAm) is estimated from DNA methylation levels at 46 CpG sites and was developed to estimate an individual’s *rate* of biological aging, expressed in years of epigenetic aging per chronological year. DunedinPoAm is based on a composite estimate of change in 18 biomarkers of organ-system integrity assessed over a 12-year period in the Dunedin cohort study [35].

### 4.5. Covariates

Several variables that might affect epigenetic aging estimates were evaluated as covariates in the main analyses based on previous research [34,42,50,61,62,98], including age, biological sex (male as the reference group), and race/ethnicity (non-Hispanic Black, Hispanic, non-Hispanic other race, and non-Hispanic white as the reference group). Secondary analyses also considered educational attainment (less than high school, high school diploma or GED, some college, and a college diploma or higher as the reference group), BMI category (25 to < 30 as overweight, 30 to < 35 as obese I, ≥ 35 as obese II, and < 25 as normal or underweight as the reference group), smoking status (current, past, and never as the reference group), and alcohol use (1-4 drinks per day, 5+ drinks per day, and none as the reference group).

### 4.6. Data analysis plan

We conducted a set of generalized linear models (GLMs) by regressing the epigenetic aging measures on each support or strain measure and adjusting for age, sex, and race/ethnicity as covariates. We then performed an additional set of models as secondary analyses that further adjusted for educational attainment and lifestyle factors, including BMI category, smoking status, and alcohol use. For both sets of analyses, we applied a 5% false discovery rate correction for multiple testing [99] across the five epigenetic aging measures for each social support and strain domain. Observations were weighted to be nationally representative of older U.S. adults using sampling weights provided by HRS. For participants who did not have specific weights for the Venous Blood Sample, weights from the 2016 core interview were used. All analyses were conducted in R 4.1.3 “One Push-Up” using the tidyverse, jtools, and survey packages [100–103].

## Data Availability

All data produced are available online at: https://hrs.isr.umich.edu/data-products

https://hrs.isr.umich.edu/data-products

## Author Contributions

Kelly E. Rentscher: Conceptualization, Writing–Original Draft; Eric T. Klopack: Conceptualization, Formal analysis, Writing–Review & Editing, Eileen M. Crimmins: Conceptualization, Writing–Review & Editing, Teresa E. Seeman: Writing–Review & Editing, Steve W. Cole: Writing–Review & Editing; Judith E. Carroll: Conceptualization, Writing– Review & Editing.

## Acknowledgements

The Health and Retirement Study is sponsored by the National Institute on Aging [U01AG009740] and is conducted by the University of Michigan.

## Supplemental Information

Table 1. Generalized linear models with spousal support predicting epigenetic aging.

Table 2. Generalized linear models with spousal strain predicting epigenetic aging.

Table 3. Generalized linear models with support from children predicting epigenetic aging.

Table 4. Generalized linear models with strain with children predicting epigenetic aging.

Table 5. Generalized linear models with support from other family members predicting epigenetic aging.

Table 6. Generalized linear models with strain with other family members predicting epigenetic aging.

Table 7. Generalized linear models with support from friends predicting epigenetic aging.

Table 8. Generalized linear models with strain with friends predicting epigenetic aging.

