## Supplemental Information for "Lower Social Support is Associated with Accelerated Epigenetic Aging: Results from the Health and Retirement Study"

Table 1

Generalized linear models with spousal support predicting epigenetic aging.

| | Horvath | | | Hannum | | | PhenoAge | | | GrimAge | | | Dunedin PoAm | | |
|---|---|---|---|---|---|---|---|---|---|---|---|---|---|---|---|
| Models | *B* | *SE* | *95% CI* | *B* | *SE* | *95% CI* | *B* | *SE* | *95% CI* | *B* | *SE* | *95% CI* | *B* | *SE* | *95% CI* |
| 1. Support with spouse | 0.03 | 0.23 | -0.43, 0.49 | -0.38 | 0.18 | -0.75, -0.01 | **-0.61** | **0.25** | **-1.11, -0.11** | **-0.34** | **0.12** | **-0.59, -0.09** | **-0.008** | **0.003** | **-0.015, -0.002** |
| Age | 0.76 | 0.02 | 0.72, 0.79 | 0.80 | 0.01 | 0.77, 0.82 | 0.79 | 0.02 | 0.74, 0.83 | 0.78 | 0.01 | 0.76, 0.80 | 0.001 | 0.000 | 0.000, 0.001 |
| Female sex | -0.69 | 0.28 | -1.25, -0.13 | -1.82 | 0.24 | -2.30, -1.33 | -1.12 | 0.38 | -1.89, -0.35 | -3.25 | 0.24 | -3.73, -2.77 | -0.020 | 0.004 | -0.029, -0.011 |
| Race/ethnicity | | | | | | | | | | | | | | | |
| Non-Hispanic Black | 0.27 | 0.50 | -0.73, 1.27 | -1.52 | 0.44 | -2.40, -0.64 | 0.65 | 0.69 | -0.74, 2.04 | 1.68 | 0.34 | 1.00, 2.37 | 0.028 | 0.009 | 0.011, 0.046 |
| Hispanic | -1.33 | 0.55 | -2.43, -0.23 | 0.06 | 0.36 | -0.65, 0.77 | -0.06 | 0.39 | -0.85, 0.72 | 0.17 | 0.36 | -0.55, 0.89 | 0.009 | 0.008 | -0.007, 0.025 |
| Non-Hispanic other race | 0.85 | 0.65 | -0.47, 2.16 | -0.30 | 0.52 | -1.35, 0.74 | -0.47 | 1.03 | -2.53, 1.60 | -0.08 | 0.60 | -1.28, 1.13 | 0.021 | 0.015 | -0.010, 0.052 |
| *N* = 2,714 | | | | | | | | | | | | | | | |
| 2. Support with spouse | 0.03 | 0.22 | -0.42, 0.47 | -0.34 | 0.17 | -0.69, 0.01 | -0.53 | 0.25 | -1.03, -0.03 | -0.10 | 0.12 | -0.35, 0.15 | -0.005 | 0.003 | -0.011, 0.002 |
| Age | 0.77 | 0.02 | 0.73, 0.81 | 0.80 | 0.01 | 0.78, 0.83 | 0.80 | 0.02 | 0.75, 0.84 | 0.81 | 0.01 | 0.79, 0.83 | 0.001 | 0.000 | 0.001, 0.002 |
| Female sex | -0.56 | 0.26 | -1.08, -0.04 | -1.77 | 0.25 | -2.28, -1.26 | -1.00 | 0.41 | -1.82, -0.17 | -2.78 | 0.21 | -3.20, -2.37 | -0.013 | 0.004 | -0.021, -0.006 |
| Race/ethnicity | | | | | | | | | | | | | | | |
| Non-Hispanic Black | 0.31 | 0.51 | -0.72, 1.33 | -1.64 | 0.47 | -2.58, -0.69 | 0.44 | 0.73 | -1.03, 1.92 | 1.19 | 0.33 | 0.52, 1.87 | 0.021 | 0.008 | 0.005, 0.038 |
| Hispanic | -1.22 | 0.53 | -2.29, -0.15 | -0.10 | 0.40 | -0.91, 0.70 | -0.54 | 0.46 | -1.46, 0.38 | -0.47 | 0.33 | -1.13, 0.20 | 0.001 | 0.008 | -0.016, 0.018 |
| Non-Hispanic other race | 0.96 | 0.65 | -0.37, 2.28 | -0.33 | 0.54 | -1.42, 0.76 | -0.40 | 0.94 | -2.31, 1.51 | 0.26 | 0.57 | -0.90, 1.41 | 0.026 | 0.015 | -0.005, 0.056 |
| Educational attainment | | | | | | | | | | | | | | | |
| Less than high school | -0.63 | 0.42 | -1.48, 0.22 | 0.02 | 0.44 | -0.86, 0.90 | 0.99 | 0.64 | -0.29, 2.28 | 1.73 | 0.27 | 1.19, 2.28 | 0.020 | 0.006 | 0.008, 0.032 |
| High school diploma/GED | -0.23 | 0.36 | -0.97, 0.51 | 0.04 | 0.29 | -0.54, 0.62 | 0.25 | 0.46 | -0.68, 1.18 | 1.15 | 0.23 | 0.69, 1.62 | 0.011 | 0.005 | 0.002, 0.021 |
| Some college | -0.41 | 0.41 | -1.23, 0.42 | -0.30 | 0.34 | -0.99, 0.40 | 0.25 | 0.40 | -0.55, 1.06 | 0.94 | 0.27 | 0.41, 1.48 | 0.009 | 0.005 | -0.002, 0.019 |
| BMI category | | | | | | | | | | | | | | | |
| Overweight (≥ 25 to < 30) | -0.35 | 0.38 | -1.11, 0.42 | -0.19 | 0.34 | -0.87, 0.49 | -0.32 | 0.46 | -1.24, 0.61 | 0.13 | 0.24 | -0.36, 0.61 | -0.001 | 0.005 | -0.010, 0.009 |
| Obese I (≥ 30 to < 35) | 0.43 | 0.42 | -0.41, 1.28 | 0.41 | 0.36 | -0.31, 1.13 | 0.48 | 0.44 | -0.42, 1.38 | 0.80 | 0.28 | 0.24, 1.37 | 0.009 | 0.004 | 0.000, 0.018 |
| Obese II (≥ 35) | 1.04 | 0.54 | -0.04, 2.13 | 1.11 | 0.37 | 0.36, 1.87 | 1.39 | 0.59 | 0.20, 2.57 | 1.76 | 0.26 | 1.23, 2.28 | 0.032 | 0.006 | 0.020, 0.045 |
| Smoking status | | | | | | | | | | | | | | | |
| Current | 0.01 | 0.59 | -1.18, 1.20 | 0.58 | 0.43 | -0.29, 1.44 | 1.16 | 0.63 | -0.11, 2.43 | 7.25 | 0.32 | 6.60, 7.89 | 0.125 | 0.008 | 0.108, 0.142 |
| Former | 0.38 | 0.31 | -0.23, 1.00 | 0.11 | 0.26 | -0.41, 0.64 | 0.26 | 0.30 | -0.34, 0.87 | 1.94 | 0.16 | 1.62, 2.26 | 0.027 | 0.004 | 0.020, 0.035 |
| Alcohol use | | | | | | | | | | | | | | | |
| 1-4 drinks per day | -0.13 | 0.34 | -0.82, 0.55 | -0.32 | 0.25 | -0.84, 0.19 | -0.05 | 0.39 | -0.83, 0.74 | -0.24 | 0.16 | -0.57, 0.09 | -0.004 | 0.004 | -0.011, 0.004 |
| 5+ drinks per day | 1.86 | 1.11 | -0.39, 4.10 | 0.85 | 0.75 | -0.66, 2.36 | 2.38 | 0.85 | 0.65, 4.10 | 1.51 | 0.72 | 0.07, 2.96 | 0.005 | 0.013 | -0.021, 0.031 |
| *N* = 2,714 | | | | | | | | | | | | | | | |

Note. PoAm = Pace of Aging methylation; *CI* = confidence interval. Bold font denotes statistically significant association following false discovery rate correction.

Table 2

Generalized linear models with spousal strain predicting epigenetic aging.

| | Horvath | | | Hannum | | | PhenoAge | | | GrimAge | | | Dunedin PoAm | | |
|---|---|---|---|---|---|---|---|---|---|---|---|---|---|---|---|
| Models | *B* | *SE* | *95% CI* | *B* | *SE* | *95% CI* | *B* | *SE* | *95% CI* | *B* | *SE* | *95% CI* | *B* | *SE* | *95% CI* |
| 1. Strain with spouse | 0.01 | 0.22 | -0.43, 0.46 | 0.35 | 0.14 | 0.06, 0.63 | 0.17 | 0.21 | -0.26, 0.60 | -0.15 | 0.14 | -0.42, 0.13 | 0.003 | 0.003 | -0.003, 0.010 |
| Age | 0.76 | 0.02 | 0.72, 0.80 | 0.80 | 0.01 | 0.77, 0.83 | 0.79 | 0.02 | 0.75, 0.83 | 0.78 | 0.01 | 0.76, 0.80 | 0.001 | 0.000 | 0.000, 0.001 |
| Female sex | -0.69 | 0.28 | -1.24, -0.13 | -1.73 | 0.24 | -2.21, -1.26 | -1.01 | 0.38 | -1.78, -0.24 | -3.21 | 0.23 | -3.67, -2.74 | -0.018 | 0.005 | -0.027, -0.009 |
| Race/ethnicity | | | | | | | | | | | | | | | |
| Non-Hispanic Black | 0.22 | 0.52 | -0.82, 1.26 | -1.42 | 0.43 | -2.29, -0.55 | 0.84 | 0.71 | -0.59, 2.28 | 1.75 | 0.35 | 1.05, 2.45 | 0.030 | 0.008 | 0.013, 0.047 |
| Hispanic | -1.38 | 0.55 | -2.49, -0.28 | 0.12 | 0.35 | -0.59, 0.83 | 0.06 | 0.39 | -0.72, 0.84 | 0.27 | 0.36 | -0.44, 0.99 | 0.010 | 0.008 | -0.006, 0.026 |
| Non-Hispanic other race | 0.81 | 0.65 | -0.50, 2.11 | -0.36 | 0.52 | -1.40, 0.69 | -0.34 | 1.01 | -2.37, 1.68 | 0.05 | 0.59 | -1.13, 1.23 | 0.023 | 0.015 | -0.008, 0.054 |
| *N* = 2,699 | | | | | | | | | | | | | | | |
| 2. Strain with spouse | -0.03 | 0.22 | -0.47, 0.42 | 0.30 | 0.14 | 0.02, 0.57 | 0.11 | 0.22 | -0.33, 0.55 | -0.21 | 0.12 | -0.44, 0.03 | 0.002 | 0.003 | -0.004, 0.009 |
| Age | 0.77 | 0.02 | 0.73, 0.81 | 0.81 | 0.01 | 0.78, 0.83 | 0.80 | 0.02 | 0.76, 0.85 | 0.81 | 0.01 | 0.79, 0.83 | 0.001 | 0.000 | 0.001, 0.002 |
| Female sex | -0.55 | 0.26 | -1.07, -0.02 | -1.69 | 0.24 | -2.18, -1.20 | -0.89 | 0.40 | -1.70, -0.08 | -2.78 | 0.20 | -3.19, -2.37 | -0.013 | 0.004 | -0.021, -0.005 |
| Race/ethnicity | | | | | | | | | | | | | | | |
| Non-Hispanic Black | 0.27 | 0.52 | -0.79, 1.33 | -1.55 | 0.46 | -2.47, -0.62 | 0.60 | 0.76 | -0.93, 2.14 | 1.18 | 0.35 | 0.48, 1.88 | 0.022 | 0.008 | 0.006, 0.039 |
| Hispanic | -1.24 | 0.53 | -2.32, -0.17 | -0.07 | 0.38 | -0.84, 0.71 | -0.46 | 0.46 | -1.39, 0.48 | -0.46 | 0.34 | -1.14, 0.23 | 0.001 | 0.009 | -0.016, 0.019 |
| Non-Hispanic other race | 0.93 | 0.65 | -0.39, 2.25 | -0.39 | 0.54 | -1.48, 0.71 | -0.29 | 0.92 | -2.16, 1.57 | 0.29 | 0.55 | -0.83, 1.41 | 0.026 | 0.015 | -0.004, 0.057 |
| Educational attainment | | | | | | | | | | | | | | | |
| Less than high school | -0.66 | 0.44 | -1.54, 0.22 | 0.07 | 0.43 | -0.80, 0.93 | 1.09 | 0.66 | -0.24, 2.42 | 1.85 | 0.25 | 1.34, 2.36 | 0.021 | 0.006 | 0.009, 0.033 |
| High school diploma/GED | -0.23 | 0.37 | -0.96, 0.51 | 0.02 | 0.28 | -0.54, 0.59 | 0.29 | 0.47 | -0.66, 1.23 | 1.17 | 0.24 | 0.69, 1.64 | 0.012 | 0.005 | 0.002, 0.021 |
| Some college | -0.37 | 0.41 | -1.21, 0.47 | -0.28 | 0.34 | -0.97, 0.41 | 0.31 | 0.40 | -0.49, 1.11 | 0.96 | 0.26 | 0.44, 1.48 | 0.009 | 0.005 | -0.001, 0.020 |
| BMI category | | | | | | | | | | | | | | | |
| Overweight (≥ 25 to < 30) | -0.33 | 0.38 | -1.09, 0.44 | -0.19 | 0.34 | -0.88, 0.51 | -0.29 | 0.47 | -1.24, 0.66 | 0.15 | 0.23 | -0.33, 0.62 | 0.000 | 0.005 | -0.010, 0.009 |
| Obese I (≥ 30 to < 35) | 0.46 | 0.42 | -0.39, 1.31 | 0.40 | 0.37 | -0.34, 1.14 | 0.49 | 0.44 | -0.41, 1.39 | 0.81 | 0.28 | 0.24, 1.37 | 0.009 | 0.004 | 0.000, 0.018 |
| Obese II (≥ 35) | 1.13 | 0.55 | 0.02, 2.24 | 1.13 | 0.37 | 0.39, 1.88 | 1.48 | 0.58 | 0.32, 2.64 | 1.81 | 0.27 | 1.27, 2.36 | 0.032 | 0.006 | 0.019, 0.045 |
| Smoking status | | | | | | | | | | | | | | | |
| Current | 0.05 | 0.59 | -1.15, 1.25 | 0.61 | 0.43 | -0.26, 1.48 | 1.21 | 0.63 | -0.06, 2.48 | 7.22 | 0.31 | 6.59, 7.86 | 0.125 | 0.008 | 0.109, 0.142 |
| Former | 0.40 | 0.31 | -0.23, 1.02 | 0.16 | 0.27 | -0.38, 0.70 | 0.31 | 0.29 | -0.28, 0.90 | 1.95 | 0.16 | 1.64, 2.27 | 0.028 | 0.004 | 0.020, 0.035 |
| Alcohol use | | | | | | | | | | | | | | | |
| 1-4 drinks per day | -0.13 | 0.34 | -0.81, 0.55 | -0.37 | 0.26 | -0.89, 0.16 | -0.07 | 0.39 | -0.86, 0.73 | -0.27 | 0.16 | -0.60, 0.06 | -0.004 | 0.004 | -0.011, 0.003 |
| 5+ drinks per day | 1.87 | 1.10 | -0.36, 4.10 | 0.80 | 0.76 | -0.74, 2.34 | 2.30 | 0.86 | 0.56, 4.03 | 1.45 | 0.71 | 0.01, 2.89 | 0.004 | 0.013 | -0.022, 0.030 |
| *N* = 2,699 | | | | | | | | | | | | | | | |

Note. PoAm = Pace of Aging methylation; *CI* = confidence interval. Bold font denotes statistically significant association following false discovery rate correction.

Table 3

Generalized linear models with support with children predicting epigenetic aging.

| | Horvath | | | Hannum | | | PhenoAge | | | GrimAge | | | Dunedin PoAm | | |
|---|---|---|---|---|---|---|---|---|---|---|---|---|---|---|---|
| Models | *B* | *SE* | *95% CI* | *B* | *SE* | *95% CI* | *B* | *SE* | *95% CI* | *B* | *SE* | *95% CI* | *B* | *SE* | *95% CI* |
| 1. Support with children | -0.16 | 0.21 | -0.59, 0.27 | -0.15 | 0.16 | -0.46, 0.17 | **-0.47** | **0.20** | **-0.87, -0.06** | **-0.39** | **0.12** | **-0.63, -0.15** | **-0.008** | **0.003** | **-0.013, -0.003** |
| Age | 0.75 | 0.02 | 0.72, 0.78 | 0.79 | 0.01 | 0.77, 0.81 | 0.78 | 0.02 | 0.75, 0.82 | 0.78 | 0.01 | 0.76, 0.80 | 0.001 | 0.000 | 0.001, 0.001 |
| Female sex | -0.92 | 0.26 | -1.44, -0.39 | -1.83 | 0.21 | -2.25, -1.42 | -1.23 | 0.35 | -1.93, -0.53 | -3.18 | 0.21 | -3.60, -2.75 | -0.018 | 0.004 | -0.026, -0.009 |
| Race/ethnicity | | | | | | | | | | | | | | | |
| Non-Hispanic Black | 0.04 | 0.40 | -0.77, 0.84 | -1.67 | 0.35 | -2.38, -0.96 | 0.71 | 0.52 | -0.33, 1.76 | 1.84 | 0.27 | 1.29, 2.40 | 0.034 | 0.007 | 0.020, 0.048 |
| Hispanic | -1.38 | 0.44 | -2.26, -0.50 | 0.19 | 0.27 | -0.36, 0.74 | 0.06 | 0.33 | -0.61, 0.73 | 0.14 | 0.30 | -0.47, 0.75 | 0.010 | 0.007 | -0.005, 0.024 |
| Non-Hispanic other race | 0.93 | 0.58 | -0.24, 2.10 | -0.19 | 0.47 | -1.14, 0.75 | 0.58 | 1.00 | -1.42, 2.58 | 0.25 | 0.53 | -0.81, 1.32 | 0.024 | 0.014 | -0.004, 0.053 |
| *N* = 3,306 | | | | | | | | | | | | | | | |
| 2. Support with children | -0.13 | 0.22 | -0.56, 0.31 | -0.10 | 0.16 | -0.42, 0.22 | -0.39 | 0.20 | -0.80, 0.02 | -0.25 | 0.11 | -0.46, -0.03 | -0.006 | 0.002 | -0.010, -0.001 |
| Age | 0.76 | 0.02 | 0.72, 0.79 | 0.79 | 0.01 | 0.77, 0.82 | 0.79 | 0.02 | 0.76, 0.83 | 0.80 | 0.01 | 0.78, 0.82 | 0.001 | 0.000 | 0.001, 0.002 |
| Female sex | -0.89 | 0.26 | -1.42, -0.36 | -1.89 | 0.21 | -2.32, -1.47 | -1.20 | 0.37 | -1.94, -0.45 | -2.97 | 0.18 | -3.33, -2.60 | -0.016 | 0.004 | -0.024, -0.009 |
| Race/ethnicity | | | | | | | | | | | | | | | |
| Non-Hispanic Black | -0.01 | 0.41 | -0.84, 0.81 | -1.85 | 0.38 | -2.61, -1.09 | 0.45 | 0.56 | -0.69, 1.59 | 1.35 | 0.27 | 0.80, 1.90 | 0.026 | 0.007 | 0.012, 0.040 |
| Hispanic | -1.38 | 0.44 | -2.26, -0.49 | -0.02 | 0.31 | -0.65, 0.61 | -0.45 | 0.45 | -1.36, 0.45 | -0.49 | 0.29 | -1.09, 0.10 | 0.001 | 0.007 | -0.014, 0.016 |
| Non-Hispanic other race | 0.97 | 0.58 | -0.20, 2.15 | -0.28 | 0.50 | -1.28, 0.72 | 0.61 | 0.93 | -1.26, 2.48 | 0.57 | 0.51 | -0.46, 1.60 | 0.029 | 0.014 | 0.000, 0.057 |
| Educational attainment | | | | | | | | | | | | | | | |
| Less than high school | -0.18 | 0.45 | -1.09, 0.72 | 0.13 | 0.43 | -0.74, 1.00 | 1.08 | 0.62 | -0.18, 2.34 | 1.76 | 0.22 | 1.31, 2.21 | 0.022 | 0.005 | 0.012, 0.032 |
| High school diploma/GED | -0.14 | 0.35 | -0.84, 0.56 | -0.01 | 0.26 | -0.54, 0.52 | 0.27 | 0.41 | -0.55, 1.09 | 1.16 | 0.17 | 0.82, 1.51 | 0.014 | 0.005 | 0.005, 0.024 |
| Some college | -0.32 | 0.34 | -1.00, 0.37 | -0.41 | 0.30 | -1.02, 0.21 | 0.27 | 0.40 | -0.53, 1.08 | 1.01 | 0.21 | 0.59, 1.42 | 0.011 | 0.004 | 0.003, 0.019 |
| BMI category | | | | | | | | | | | | | | | |
| Overweight (≥ 25 to < 30) | -0.31 | 0.35 | -1.02, 0.40 | -0.16 | 0.33 | -0.84, 0.51 | -0.21 | 0.41 | -1.03, 0.62 | 0.08 | 0.23 | -0.38, 0.55 | -0.001 | 0.004 | -0.010, 0.008 |
| Obese I (≥ 30 to < 35) | 0.35 | 0.39 | -0.42, 1.13 | 0.29 | 0.36 | -0.45, 1.02 | 0.41 | 0.43 | -0.45, 1.27 | 0.52 | 0.27 | -0.02, 1.06 | 0.009 | 0.004 | 0.001, 0.017 |
| Obese II (≥ 35) | 0.99 | 0.51 | -0.03, 2.01 | 1.09 | 0.38 | 0.32, 1.86 | 1.64 | 0.62 | 0.39, 2.88 | 1.54 | 0.27 | 1.00, 2.07 | 0.032 | 0.005 | 0.022, 0.043 |
| Smoking status | | | | | | | | | | | | | | | |
| Current | 0.17 | 0.52 | -0.88, 1.22 | 0.58 | 0.38 | -0.19, 1.35 | 1.11 | 0.55 | -0.01, 2.23 | 7.46 | 0.32 | 6.82, 8.10 | 0.126 | 0.007 | 0.111, 0.140 |
| Former | 0.24 | 0.26 | -0.29, 0.77 | 0.01 | 0.23 | -0.46, 0.47 | 0.37 | 0.28 | -0.19, 0.93 | 1.99 | 0.13 | 1.72, 2.26 | 0.024 | 0.003 | 0.017, 0.031 |
| Alcohol use | | | | | | | | | | | | | | | |
| 1-4 drinks per day | -0.10 | 0.25 | -0.61, 0.40 | -0.50 | 0.22 | -0.94, -0.07 | -0.21 | 0.35 | -0.93, 0.50 | -0.38 | 0.13 | -0.64, -0.11 | -0.006 | 0.003 | -0.013, 0.000 |
| 5+ drinks per day | 1.13 | 0.99 | -0.86, 3.12 | 0.66 | 0.71 | -0.78, 2.10 | 2.56 | 0.84 | 0.87, 4.25 | 0.83 | 0.54 | -0.26, 1.92 | -0.007 | 0.011 | -0.028, 0.015 |
| *N* = 3,306 | | | | | | | | | | | | | | | |

Note. PoAm = Pace of Aging methylation; *CI* = confidence interval. Bold font denotes statistically significant association following false discovery rate correction.

Table 4

Generalized linear models with strain with children predicting epigenetic aging.

| | Horvath | | | Hannum | | | PhenoAge | | | GrimAge | | | Dunedin PoAm | | |
|---|---|---|---|---|---|---|---|---|---|---|---|---|---|---|---|
| Models | *B* | *SE* | *95% CI* | *B* | *SE* | *95% CI* | *B* | *SE* | *95% CI* | *B* | *SE* | *95% CI* | *B* | *SE* | *95% CI* |
| 1. Strain with children | 0.21 | 0.19 | -0.16, 0.59 | 0.16 | 0.15 | -0.14, 0.47 | 0.49 | 0.28 | -0.07, 1.06 | 0.32 | 0.14 | 0.05, 0.59 | 0.006 | 0.003 | 0.001, 0.011 |
| Age | 0.75 | 0.02 | 0.72, 0.78 | 0.79 | 0.01 | 0.77, 0.82 | 0.78 | 0.02 | 0.75, 0.82 | 0.78 | 0.01 | 0.76, 0.79 | 0.001 | 0.000 | 0.001, 0.001 |
| Female sex | -0.97 | 0.26 | -1.50, -0.44 | -1.88 | 0.21 | -2.30, -1.45 | -1.35 | 0.36 | -2.07, -0.62 | -3.28 | 0.21 | -3.70, -2.87 | -0.020 | 0.004 | -0.028, -0.011 |
| Race/ethnicity | | | | | | | | | | | | | | | |
| Non-Hispanic Black | 0.02 | 0.40 | -0.79, 0.83 | -1.71 | 0.34 | -2.39, -1.03 | 0.62 | 0.50 | -0.39, 1.63 | 1.81 | 0.27 | 1.26, 2.36 | 0.033 | 0.007 | 0.019, 0.047 |
| Hispanic | -1.36 | 0.44 | -2.25, -0.48 | 0.13 | 0.27 | -0.41, 0.68 | 0.04 | 0.33 | -0.63, 0.70 | 0.10 | 0.30 | -0.51, 0.71 | 0.009 | 0.007 | -0.005, 0.023 |
| Non-Hispanic other race | 0.90 | 0.59 | -0.28, 2.09 | -0.21 | 0.48 | -1.17, 0.74 | 0.51 | 0.99 | -1.48, 2.50 | 0.20 | 0.55 | -0.89, 1.30 | 0.024 | 0.015 | -0.006, 0.053 |
| *N* = 3,314 | | | | | | | | | | | | | | | |
| 2. Strain with children | 0.16 | 0.19 | -0.22, 0.55 | 0.09 | 0.14 | -0.21, 0.38 | 0.38 | 0.29 | -0.21, 0.96 | 0.06 | 0.11 | -0.16, 0.29 | 0.002 | 0.002 | -0.003, 0.007 |
| Age | 0.76 | 0.02 | 0.72, 0.79 | 0.79 | 0.01 | 0.77, 0.82 | 0.79 | 0.02 | 0.76, 0.83 | 0.80 | 0.01 | 0.78, 0.82 | 0.001 | 0.000 | 0.001, 0.002 |
| Female sex | -0.94 | 0.26 | -1.47, -0.40 | -1.92 | 0.22 | -2.36, -1.49 | -1.29 | 0.38 | -2.06, -0.53 | -3.03 | 0.18 | -3.39, -2.67 | -0.018 | 0.004 | -0.025, -0.010 |
| Race/ethnicity | | | | | | | | | | | | | | | |
| Non-Hispanic Black | -0.02 | 0.41 | -0.84, 0.81 | -1.89 | 0.36 | -2.62, -1.15 | 0.37 | 0.55 | -0.74, 1.48 | 1.34 | 0.27 | 0.79, 1.89 | 0.025 | 0.007 | 0.011, 0.039 |
| Hispanic | -1.37 | 0.44 | -2.25, -0.48 | -0.09 | 0.31 | -0.73, 0.54 | -0.47 | 0.45 | -1.37, 0.43 | -0.52 | 0.28 | -1.09, 0.06 | 0.001 | 0.007 | -0.014, 0.015 |
| Non-Hispanic other race | 0.95 | 0.59 | -0.24, 2.14 | -0.29 | 0.50 | -1.31, 0.72 | 0.55 | 0.92 | -1.31, 2.42 | 0.55 | 0.52 | -0.50, 1.60 | 0.028 | 0.014 | -0.001, 0.057 |
| Educational attainment | | | | | | | | | | | | | | | |
| Less than high school | -0.18 | 0.44 | -1.08, 0.71 | 0.16 | 0.44 | -0.72, 1.05 | 1.09 | 0.62 | -0.18, 2.35 | 1.78 | 0.22 | 1.33, 2.23 | 0.022 | 0.005 | 0.012, 0.033 |
| High school diploma/GED | -0.15 | 0.35 | -0.86, 0.55 | 0.00 | 0.27 | -0.54, 0.54 | 0.26 | 0.41 | -0.56, 1.08 | 1.17 | 0.17 | 0.83, 1.52 | 0.014 | 0.005 | 0.005, 0.024 |
| Some college | -0.33 | 0.34 | -1.01, 0.35 | -0.39 | 0.31 | -1.02, 0.23 | 0.25 | 0.40 | -0.55, 1.06 | 1.01 | 0.21 | 0.59, 1.43 | 0.011 | 0.004 | 0.003, 0.019 |
| BMI category | | | | | | | | | | | | | | | |
| Overweight (≥ 25 to < 30) | -0.31 | 0.35 | -1.01, 0.39 | -0.14 | 0.33 | -0.80, 0.52 | -0.21 | 0.40 | -1.03, 0.61 | 0.08 | 0.23 | -0.39, 0.54 | -0.001 | 0.004 | -0.010, 0.008 |
| Obese I (≥ 30 to < 35) | 0.35 | 0.39 | -0.43, 1.14 | 0.32 | 0.36 | -0.40, 1.04 | 0.44 | 0.43 | -0.42, 1.29 | 0.53 | 0.27 | -0.02, 1.08 | 0.009 | 0.004 | 0.001, 0.017 |
| Obese II (≥ 35) | 0.97 | 0.51 | -0.05, 2.00 | 1.11 | 0.37 | 0.36, 1.87 | 1.62 | 0.62 | 0.37, 2.86 | 1.54 | 0.26 | 1.00, 2.07 | 0.032 | 0.005 | 0.022, 0.042 |
| Smoking status | | | | | | | | | | | | | | | |
| Current | 0.16 | 0.52 | -0.89, 1.21 | 0.59 | 0.38 | -0.18, 1.35 | 1.11 | 0.55 | -0.01, 2.22 | 7.47 | 0.31 | 6.83, 8.10 | 0.126 | 0.007 | 0.111, 0.140 |
| Former | 0.23 | 0.27 | -0.31, 0.77 | 0.01 | 0.23 | -0.45, 0.47 | 0.36 | 0.28 | -0.20, 0.92 | 2.00 | 0.14 | 1.73, 2.28 | 0.024 | 0.004 | 0.017, 0.031 |
| Alcohol use | | | | | | | | | | | | | | | |
| 1-4 drinks per day | -0.10 | 0.25 | -0.61, 0.40 | -0.49 | 0.21 | -0.92, -0.06 | -0.20 | 0.36 | -0.91, 0.52 | -0.37 | 0.13 | -0.64, -0.11 | -0.006 | 0.003 | -0.013, 0.000 |
| 5+ drinks per day | 1.15 | 0.98 | -0.83, 3.13 | 0.68 | 0.71 | -0.75, 2.11 | 2.60 | 0.83 | 0.91, 4.28 | 0.84 | 0.55 | -0.27, 1.94 | -0.007 | 0.010 | -0.028, 0.014 |
| *N* = 3,314 | | | | | | | | | | | | | | | |

Note. PoAm = Pace of Aging methylation; *CI* = confidence interval. Bold font denotes statistically significant association following false discovery rate correction.

Table 5

Generalized linear models with support with other family members predicting epigenetic aging.

| | Horvath | | | Hannum | | | PhenoAge | | | GrimAge | | | Dunedin PoAm | | |
|---|---|---|---|---|---|---|---|---|---|---|---|---|---|---|---|
| Models | *B* | *SE* | *95% CI* | *B* | *SE* | *95% CI* | *B* | *SE* | *95% CI* | *B* | *SE* | *95% CI* | *B* | *SE* | *95% CI* |
| 1. Support with other family | 0.03 | 0.14 | -0.25, 0.31 | -0.16 | 0.12 | -0.40, 0.07 | 0.00 | 0.14 | -0.27, 0.27 | -0.24 | 0.11 | -0.46, -0.01 | **-0.006** | **0.002** | **-0.010, -0.002** |
| Age | 0.74 | 0.02 | 0.70, 0.77 | 0.78 | 0.01 | 0.76, 0.81 | 0.78 | 0.02 | 0.74, 0.81 | 0.77 | 0.01 | 0.76, 0.79 | 0.001 | 0.000 | 0.000, 0.001 |
| Female sex | -0.89 | 0.27 | -1.42, -0.36 | -1.76 | 0.20 | -2.16, -1.36 | -1.22 | 0.31 | -1.84, -0.59 | -3.25 | 0.21 | -3.68, -2.82 | -0.017 | 0.004 | -0.025, -0.009 |
| Race/ethnicity | | | | | | | | | | | | | | | |
| Non-Hispanic Black | -0.10 | 0.38 | -0.86, 0.66 | -1.71 | 0.32 | -2.36, -1.06 | 0.83 | 0.48 | -0.13, 1.80 | 1.85 | 0.25 | 1.35, 2.35 | 0.034 | 0.007 | 0.021, 0.048 |
| Hispanic | -1.45 | 0.45 | -2.35, -0.56 | 0.07 | 0.27 | -0.47, 0.61 | 0.10 | 0.36 | -0.62, 0.83 | 0.12 | 0.30 | -0.48, 0.73 | 0.007 | 0.007 | -0.006, 0.021 |
| Non-Hispanic other race | 0.56 | 0.55 | -0.54, 1.66 | -0.28 | 0.50 | -1.28, 0.72 | 0.35 | 0.91 | -1.48, 2.17 | 0.33 | 0.49 | -0.66, 1.32 | 0.030 | 0.013 | 0.003, 0.056 |
| *N* = 3,570 | | | | | | | | | | | | | | | |
| 2. Support with other family | 0.07 | 0.14 | -0.21, 0.34 | -0.13 | 0.11 | -0.36, 0.11 | 0.06 | 0.14 | -0.22, 0.34 | -0.19 | 0.08 | -0.35, -0.02 | **-0.005** | **0.002** | **-0.008, -0.002** |
| Age | 0.74 | 0.02 | 0.71, 0.78 | 0.79 | 0.01 | 0.77, 0.81 | 0.78 | 0.02 | 0.75, 0.82 | 0.80 | 0.01 | 0.78, 0.81 | 0.001 | 0.000 | 0.001, 0.002 |
| Female sex | -0.91 | 0.26 | -1.43, -0.40 | -1.84 | 0.20 | -2.24, -1.43 | -1.22 | 0.33 | -1.88, -0.56 | -2.96 | 0.19 | -3.35, -2.57 | -0.014 | 0.003 | -0.020, -0.007 |
| Race/ethnicity | | | | | | | | | | | | | | | |
| Non-Hispanic Black | -0.14 | 0.38 | -0.91, 0.63 | -1.91 | 0.35 | -2.61, -1.21 | 0.53 | 0.52 | -0.52, 1.57 | 1.28 | 0.23 | 0.82, 1.74 | 0.025 | 0.006 | 0.013, 0.038 |
| Hispanic | -1.41 | 0.43 | -2.28, -0.53 | -0.15 | 0.33 | -0.81, 0.51 | -0.43 | 0.45 | -1.34, 0.47 | -0.46 | 0.28 | -1.02, 0.10 | 0.001 | 0.007 | -0.014, 0.015 |
| Non-Hispanic other race | 0.66 | 0.55 | -0.44, 1.77 | -0.33 | 0.52 | -1.38, 0.73 | 0.41 | 0.85 | -1.31, 2.12 | 0.58 | 0.45 | -0.34, 1.49 | 0.033 | 0.013 | 0.007, 0.059 |
| Educational attainment | | | | | | | | | | | | | | | |
| Less than high school | -0.21 | 0.42 | -1.05, 0.63 | 0.27 | 0.40 | -0.54, 1.09 | 1.29 | 0.55 | 0.17, 2.40 | 1.77 | 0.25 | 1.26, 2.27 | 0.018 | 0.005 | 0.008, 0.028 |
| High school diploma/GED | 0.10 | 0.34 | -0.58, 0.79 | 0.22 | 0.27 | -0.33, 0.78 | 0.68 | 0.42 | -0.17, 1.53 | 1.31 | 0.18 | 0.94, 1.68 | 0.014 | 0.005 | 0.005, 0.023 |
| Some college | -0.07 | 0.30 | -0.67, 0.54 | -0.09 | 0.27 | -0.64, 0.46 | 0.68 | 0.38 | -0.10, 1.46 | 1.13 | 0.22 | 0.68, 1.58 | 0.011 | 0.004 | 0.002, 0.020 |
| BMI category | | | | | | | | | | | | | | | |
| Overweight (≥ 25 to < 30) | -0.58 | 0.35 | -1.28, 0.12 | -0.11 | 0.31 | -0.74, 0.51 | -0.29 | 0.42 | -1.13, 0.56 | 0.07 | 0.20 | -0.34, 0.48 | 0.001 | 0.004 | -0.007, 0.010 |
| Obese I (≥ 30 to < 35) | 0.07 | 0.41 | -0.76, 0.89 | 0.22 | 0.35 | -0.49, 0.93 | 0.31 | 0.44 | -0.57, 1.20 | 0.48 | 0.23 | 0.03, 0.94 | 0.010 | 0.004 | 0.003, 0.018 |
| Obese II (≥ 35) | 0.58 | 0.50 | -0.43, 1.59 | 1.02 | 0.35 | 0.31, 1.73 | 1.67 | 0.52 | 0.61, 2.73 | 1.53 | 0.24 | 1.05, 2.01 | 0.033 | 0.005 | 0.023, 0.043 |
| Smoking status | | | | | | | | | | | | | | | |
| Current | 0.25 | 0.51 | -0.78, 1.29 | 0.69 | 0.34 | 0.01, 1.37 | 0.99 | 0.46 | 0.07, 1.92 | 7.48 | 0.31 | 6.85, 8.10 | 0.123 | 0.007 | 0.110, 0.137 |
| Former | 0.27 | 0.28 | -0.30, 0.84 | -0.03 | 0.24 | -0.51, 0.45 | 0.23 | 0.28 | -0.33, 0.79 | 1.91 | 0.14 | 1.62, 2.19 | 0.025 | 0.003 | 0.018, 0.031 |
| Alcohol use | | | | | | | | | | | | | | | |
| 1-4 drinks per day | -0.15 | 0.27 | -0.69, 0.38 | -0.40 | 0.21 | -0.83, 0.02 | -0.13 | 0.33 | -0.81, 0.54 | -0.35 | 0.13 | -0.62, -0.07 | -0.006 | 0.003 | -0.013, 0.001 |
| 5+ drinks per day | 1.10 | 0.81 | -0.54, 2.74 | 0.37 | 0.64 | -0.92, 1.66 | 2.35 | 0.78 | 0.78, 3.92 | 1.37 | 0.53 | 0.30, 2.44 | 0.004 | 0.011 | -0.018, 0.026 |
| *N* = 3,570 | | | | | | | | | | | | | | | |

Note. PoAm = Pace of Aging methylation; *CI* = confidence interval. Bold font denotes statistically significant association following false discovery rate correction.

Table 6

Generalized linear models with strain with other family members predicting epigenetic aging.

| | Horvath | | | Hannum | | | PhenoAge | | | GrimAge | | | Dunedin PoAm | | |
|---|---|---|---|---|---|---|---|---|---|---|---|---|---|---|---|
| Models | *B* | *SE* | *95% CI* | *B* | *SE* | *95% CI* | *B* | *SE* | *95% CI* | *B* | *SE* | *95% CI* | *B* | *SE* | *95% CI* |
| 1. Strain with other family | 0.14 | 0.24 | -0.35, 0.62 | 0.21 | 0.17 | -0.13, 0.54 | -0.01 | 0.24 | -0.50, 0.48 | 0.22 | 0.15 | -0.08, 0.52 | 0.005 | 0.003 | -0.002, 0.011 |
| Age | 0.74 | 0.02 | 0.71, 0.78 | 0.79 | 0.01 | 0.76, 0.81 | 0.78 | 0.02 | 0.74, 0.81 | 0.77 | 0.01 | 0.76, 0.79 | 0.001 | 0.000 | 0.000, 0.001 |
| Female sex | -0.91 | 0.26 | -1.44, -0.38 | -1.82 | 0.21 | -2.23, -1.40 | -1.21 | 0.32 | -1.86, -0.57 | -3.31 | 0.21 | -3.74, -2.88 | -0.019 | 0.004 | -0.027, -0.011 |
| Race/ethnicity | | | | | | | | | | | | | | | |
| Non-Hispanic Black | -0.12 | 0.39 | -0.90, 0.65 | -1.78 | 0.33 | -2.44, -1.13 | 0.84 | 0.48 | -0.13, 1.80 | 1.77 | 0.25 | 1.27, 2.27 | 0.032 | 0.007 | 0.019, 0.046 |
| Hispanic | -1.46 | 0.45 | -2.37, -0.55 | 0.01 | 0.26 | -0.51, 0.53 | 0.10 | 0.36 | -0.62, 0.82 | 0.05 | 0.30 | -0.55, 0.64 | 0.005 | 0.007 | -0.009, 0.019 |
| Non-Hispanic other race | 0.56 | 0.55 | -0.55, 1.66 | -0.31 | 0.50 | -1.31, 0.69 | 0.35 | 0.91 | -1.48, 2.18 | 0.30 | 0.50 | -0.71, 1.30 | 0.028 | 0.013 | 0.001, 0.055 |
| *N* = 3,569 | | | | | | | | | | | | | | | |
| 2. Strain with other family | 0.10 | 0.24 | -0.38, 0.58 | 0.15 | 0.17 | -0.19, 0.49 | -0.11 | 0.26 | -0.63, 0.41 | 0.02 | 0.12 | -0.23, 0.27 | 0.001 | 0.003 | -0.005, 0.007 |
| Age | 0.74 | 0.02 | 0.71, 0.78 | 0.79 | 0.01 | 0.77, 0.81 | 0.78 | 0.02 | 0.75, 0.82 | 0.80 | 0.01 | 0.78, 0.81 | 0.001 | 0.000 | 0.001, 0.002 |
| Female sex | -0.91 | 0.25 | -1.42, -0.40 | -1.88 | 0.21 | -2.30, -1.46 | -1.20 | 0.33 | -1.87, -0.53 | -3.00 | 0.19 | -3.38, -2.61 | -0.015 | 0.003 | -0.022, -0.008 |
| Race/ethnicity | | | | | | | | | | | | | | | |
| Non-Hispanic Black | -0.15 | 0.39 | -0.95, 0.64 | -1.96 | 0.35 | -2.67, -1.25 | 0.56 | 0.52 | -0.48, 1.61 | 1.24 | 0.23 | 0.77, 1.71 | 0.024 | 0.006 | 0.011, 0.037 |
| Hispanic | -1.43 | 0.44 | -2.32, -0.53 | -0.19 | 0.32 | -0.83, 0.45 | -0.41 | 0.44 | -1.31, 0.49 | -0.49 | 0.27 | -1.04, 0.05 | -0.001 | 0.007 | -0.015, 0.013 |
| Non-Hispanic other race | 0.66 | 0.55 | -0.44, 1.77 | -0.35 | 0.52 | -1.41, 0.71 | 0.42 | 0.85 | -1.30, 2.14 | 0.55 | 0.46 | -0.39, 1.49 | 0.032 | 0.013 | 0.005, 0.059 |
| Educational attainment | | | | | | | | | | | | | | | |
| Less than high school | -0.16 | 0.43 | -1.03, 0.71 | 0.26 | 0.40 | -0.56, 1.08 | 1.28 | 0.56 | 0.16, 2.40 | 1.72 | 0.25 | 1.22, 2.22 | 0.018 | 0.005 | 0.008, 0.028 |
| High school diploma/GED | 0.10 | 0.34 | -0.59, 0.79 | 0.23 | 0.27 | -0.32, 0.78 | 0.67 | 0.42 | -0.18, 1.52 | 1.32 | 0.18 | 0.94, 1.69 | 0.014 | 0.005 | 0.005, 0.023 |
| Some college | -0.07 | 0.30 | -0.68, 0.54 | -0.10 | 0.27 | -0.65, 0.45 | 0.68 | 0.39 | -0.10, 1.46 | 1.13 | 0.22 | 0.67, 1.58 | 0.011 | 0.004 | 0.002, 0.020 |
| BMI category | | | | | | | | | | | | | | | |
| Overweight (≥ 25 to < 30) | -0.57 | 0.35 | -1.27, 0.14 | -0.11 | 0.31 | -0.74, 0.52 | -0.29 | 0.42 | -1.14, 0.55 | 0.06 | 0.20 | -0.35, 0.48 | 0.002 | 0.004 | -0.007, 0.010 |
| Obese I (≥ 30 to < 35) | 0.06 | 0.41 | -0.77, 0.89 | 0.23 | 0.35 | -0.48, 0.94 | 0.31 | 0.44 | -0.58, 1.19 | 0.50 | 0.23 | 0.04, 0.97 | 0.011 | 0.004 | 0.003, 0.019 |
| Obese II (≥ 35) | 0.55 | 0.49 | -0.44, 1.55 | 1.03 | 0.35 | 0.32, 1.73 | 1.67 | 0.53 | 0.61, 2.73 | 1.57 | 0.25 | 1.07, 2.07 | 0.034 | 0.005 | 0.023, 0.044 |
| Smoking status | | | | | | | | | | | | | | | |
| Current | 0.29 | 0.51 | -0.74, 1.31 | 0.67 | 0.33 | 0.00, 1.35 | 1.00 | 0.46 | 0.07, 1.92 | 7.45 | 0.30 | 6.83, 8.06 | 0.123 | 0.007 | 0.110, 0.137 |
| Former | 0.26 | 0.28 | -0.31, 0.83 | -0.03 | 0.24 | -0.51, 0.45 | 0.23 | 0.28 | -0.33, 0.78 | 1.92 | 0.14 | 1.63, 2.20 | 0.025 | 0.003 | 0.018, 0.032 |
| Alcohol use | | | | | | | | | | | | | | | |
| 1-4 drinks per day | -0.15 | 0.27 | -0.70, 0.39 | -0.40 | 0.21 | -0.83, 0.03 | -0.14 | 0.34 | -0.82, 0.54 | -0.34 | 0.13 | -0.61, -0.07 | -0.006 | 0.003 | -0.013, 0.001 |
| 5+ drinks per day | 1.09 | 0.82 | -0.55, 2.74 | 0.39 | 0.64 | -0.90, 1.68 | 2.34 | 0.78 | 0.77, 3.91 | 1.39 | 0.54 | 0.31, 2.47 | 0.004 | 0.011 | -0.017, 0.026 |
| *N* = 3,569 | | | | | | | | | | | | | | | |

Note. PoAm = Pace of Aging methylation; *CI* = confidence interval. Bold font denotes statistically significant association following false discovery rate correction.

Table 7

Generalized linear models with support with friends predicting epigenetic aging.

| | Horvath | | | Hannum | | | PhenoAge | | | GrimAge | | | Dunedin PoAm | | |
|---|---|---|---|---|---|---|---|---|---|---|---|---|---|---|---|
| Models | *B* | *SE* | *95% CI* | *B* | *SE* | *95% CI* | *B* | *SE* | *95% CI* | *B* | *SE* | *95% CI* | *B* | *SE* | *95% CI* |
| 1. Support with friends | -0.18 | 0.17 | -0.52, 0.16 | 0.03 | 0.14 | -0.25, 0.31 | -0.08 | 0.20 | -0.49, 0.33 | **-0.38** | **0.10** | **-0.59, -0.17** | **-0.005** | **0.002** | **-0.010, -0.001** |
| Age | 0.74 | 0.02 | 0.71, 0.78 | 0.79 | 0.01 | 0.76, 0.81 | 0.78 | 0.02 | 0.75, 0.81 | 0.78 | 0.01 | 0.76, 0.79 | 0.001 | 0.000 | 0.000, 0.001 |
| Female sex | -0.88 | 0.28 | -1.45, -0.31 | -1.82 | 0.23 | -2.27, -1.37 | -1.21 | 0.31 | -1.82, -0.59 | -3.20 | 0.22 | -3.64, -2.76 | -0.018 | 0.004 | -0.026, -0.010 |
| Race/ethnicity | | | | | | | | | | | | | | | |
| Non-Hispanic Black | 0.12 | 0.40 | -0.69, 0.94 | -1.67 | 0.33 | -2.33, -1.01 | 0.89 | 0.50 | -0.11, 1.89 | 1.98 | 0.24 | 1.51, 2.46 | 0.037 | 0.007 | 0.023, 0.050 |
| Hispanic | -1.45 | 0.43 | -2.31, -0.59 | 0.00 | 0.26 | -0.53, 0.52 | 0.02 | 0.36 | -0.71, 0.74 | 0.02 | 0.29 | -0.57, 0.61 | 0.006 | 0.007 | -0.008, 0.020 |
| Non-Hispanic other race | 0.44 | 0.59 | -0.75, 1.62 | -0.53 | 0.61 | -1.76, 0.69 | 0.19 | 0.95 | -1.72, 2.11 | 0.46 | 0.41 | -0.36, 1.28 | 0.032 | 0.011 | 0.010, 0.054 |
| *N* = 3,475 | | | | | | | | | | | | | | | |
| 2. Support with friends | -0.16 | 0.17 | -0.51, 0.19 | 0.09 | 0.15 | -0.20, 0.38 | 0.02 | 0.22 | -0.41, 0.46 | **-0.26** | **0.09** | **-0.45, -0.08** | -0.004 | 0.002 | -0.008, 0.001 |
| Age | 0.75 | 0.02 | 0.71, 0.79 | 0.79 | 0.01 | 0.77, 0.81 | 0.79 | 0.02 | 0.76, 0.82 | 0.80 | 0.01 | 0.79, 0.82 | 0.001 | 0.000 | 0.001, 0.002 |
| Female sex | -0.88 | 0.27 | -1.43, -0.33 | -1.89 | 0.23 | -2.36, -1.42 | -1.20 | 0.32 | -1.84, -0.56 | -2.95 | 0.19 | -3.33, -2.57 | -0.015 | 0.003 | -0.022, -0.008 |
| Race/ethnicity | | | | | | | | | | | | | | | |
| Non-Hispanic Black | 0.10 | 0.41 | -0.72, 0.92 | -1.85 | 0.35 | -2.56, -1.15 | 0.61 | 0.53 | -0.46, 1.69 | 1.44 | 0.22 | 1.00, 1.89 | 0.028 | 0.006 | 0.015, 0.040 |
| Hispanic | -1.38 | 0.43 | -2.26, -0.50 | -0.14 | 0.31 | -0.77, 0.49 | -0.42 | 0.44 | -1.31, 0.47 | -0.47 | 0.28 | -1.03, 0.09 | 0.000 | 0.007 | -0.014, 0.015 |
| Non-Hispanic other race | 0.57 | 0.58 | -0.61, 1.75 | -0.51 | 0.62 | -1.76, 0.74 | 0.37 | 0.93 | -1.52, 2.26 | 0.84 | 0.34 | 0.15, 1.54 | 0.037 | 0.011 | 0.014, 0.060 |
| Educational attainment | | | | | | | | | | | | | | | |
| Less than high school | -0.22 | 0.42 | -1.06, 0.63 | 0.10 | 0.38 | -0.68, 0.87 | 1.14 | 0.59 | -0.07, 2.34 | 1.61 | 0.21 | 1.20, 2.03 | 0.016 | 0.005 | 0.006, 0.026 |
| High school diploma/GED | 0.02 | 0.34 | -0.66, 0.71 | 0.16 | 0.26 | -0.36, 0.69 | 0.65 | 0.44 | -0.23, 1.53 | 1.27 | 0.18 | 0.91, 1.63 | 0.012 | 0.004 | 0.003, 0.021 |
| Some college | -0.18 | 0.32 | -0.83, 0.47 | -0.15 | 0.26 | -0.68, 0.39 | 0.68 | 0.38 | -0.08, 1.45 | 1.13 | 0.22 | 0.69, 1.57 | 0.010 | 0.005 | 0.001, 0.020 |
| BMI category | | | | | | | | | | | | | | | |
| Overweight (≥ 25 to < 30) | -0.62 | 0.34 | -1.30, 0.06 | -0.12 | 0.32 | -0.77, 0.52 | -0.19 | 0.42 | -1.03, 0.65 | 0.03 | 0.21 | -0.40, 0.47 | 0.002 | 0.004 | -0.007, 0.010 |
| Obese I (≥ 30 to < 35) | 0.10 | 0.40 | -0.70, 0.90 | 0.29 | 0.36 | -0.43, 1.02 | 0.21 | 0.42 | -0.63, 1.06 | 0.53 | 0.24 | 0.05, 1.00 | 0.012 | 0.004 | 0.003, 0.020 |
| Obese II (≥ 35) | 0.60 | 0.52 | -0.44, 1.65 | 1.15 | 0.39 | 0.37, 1.93 | 1.83 | 0.52 | 0.78, 2.89 | 1.57 | 0.26 | 1.05, 2.10 | 0.033 | 0.005 | 0.022, 0.044 |
| Smoking status | | | | | | | | | | | | | | | |
| Current | 0.16 | 0.49 | -0.84, 1.15 | 0.69 | 0.33 | 0.02, 1.36 | 0.89 | 0.47 | -0.07, 1.85 | 7.36 | 0.28 | 6.80, 7.93 | 0.123 | 0.007 | 0.109, 0.137 |
| Former | 0.17 | 0.28 | -0.39, 0.73 | 0.00 | 0.23 | -0.47, 0.47 | 0.25 | 0.27 | -0.31, 0.80 | 1.98 | 0.14 | 1.69, 2.27 | 0.026 | 0.003 | 0.019, 0.033 |
| Alcohol use | | | | | | | | | | | | | | | |
| 1-4 drinks per day | -0.08 | 0.26 | -0.60, 0.44 | -0.42 | 0.22 | -0.86, 0.03 | -0.08 | 0.34 | -0.77, 0.61 | -0.25 | 0.14 | -0.53, 0.04 | -0.006 | 0.004 | -0.013, 0.001 |
| 5+ drinks per day | 1.53 | 0.81 | -0.10, 3.16 | 0.52 | 0.66 | -0.82, 1.86 | 2.61 | 0.80 | 1.00, 4.22 | 1.36 | 0.54 | 0.27, 2.44 | 0.003 | 0.011 | -0.019, 0.025 |
| *N* = 3,475 | | | | | | | | | | | | | | | |

Note. PoAm = Pace of Aging methylation; *CI* = confidence interval. Bold font denotes statistically significant association following false discovery rate correction.

Table 8

Generalized linear models with strain with friends predicting epigenetic aging.

| | Horvath | | | Hannum | | | PhenoAge | | | GrimAge | | | Dunedin PoAm | | |
|---|---|---|---|---|---|---|---|---|---|---|---|---|---|---|---|
| Models | *B* | *SE* | *95% CI* | *B* | *SE* | *95% CI* | *B* | *SE* | *95% CI* | *B* | *SE* | *95% CI* | *B* | *SE* | *95% CI* |
| 1. Strain with friends | 0.04 | 0.31 | -0.58, 0.66 | 0.16 | 0.23 | -0.30, 0.62 | -0.33 | 0.35 | -1.02, 0.37 | 0.43 | 0.20 | 0.02, 0.84 | **0.013** | **0.004** | **0.005, 0.021** |
| Age | 0.75 | 0.02 | 0.71, 0.78 | 0.79 | 0.01 | 0.76, 0.81 | 0.78 | 0.02 | 0.74, 0.81 | 0.78 | 0.01 | 0.76, 0.80 | 0.001 | 0.000 | 0.001, 0.001 |
| Female sex | -0.93 | 0.27 | -1.48, -0.38 | -1.80 | 0.22 | -2.25, -1.36 | -1.25 | 0.30 | -1.85, -0.65 | -3.33 | 0.21 | -3.74, -2.91 | -0.019 | 0.004 | -0.027, -0.012 |
| Race/ethnicity | | | | | | | | | | | | | | | |
| Non-Hispanic Black | 0.12 | 0.42 | -0.72, 0.96 | -1.70 | 0.33 | -2.36, -1.03 | 0.94 | 0.49 | -0.05, 1.94 | 1.93 | 0.24 | 1.45, 2.41 | 0.035 | 0.007 | 0.021, 0.048 |
| Hispanic | -1.27 | 0.44 | -2.15, -0.40 | 0.00 | 0.27 | -0.54, 0.53 | 0.02 | 0.35 | -0.69, 0.73 | 0.09 | 0.30 | -0.52, 0.69 | 0.007 | 0.007 | -0.007, 0.021 |
| Non-Hispanic other race | 0.45 | 0.60 | -0.76, 1.66 | -0.57 | 0.61 | -1.80, 0.66 | 0.27 | 0.93 | -1.60, 2.14 | 0.42 | 0.41 | -0.41, 1.24 | 0.030 | 0.011 | 0.009, 0.051 |
| *N* = 3,473 | | | | | | | | | | | | | | | |
| 2. Strain with friends | -0.03 | 0.30 | -0.64, 0.58 | 0.04 | 0.21 | -0.40, 0.47 | -0.51 | 0.33 | -1.18, 0.16 | -0.09 | 0.16 | -0.42, 0.25 | 0.004 | 0.004 | -0.003, 0.012 |
| Age | 0.75 | 0.02 | 0.71, 0.79 | 0.79 | 0.01 | 0.77, 0.82 | 0.79 | 0.02 | 0.75, 0.82 | 0.80 | 0.01 | 0.79, 0.82 | 0.001 | 0.000 | 0.001, 0.002 |
| Female sex | -0.93 | 0.26 | -1.46, -0.39 | -1.85 | 0.23 | -2.31, -1.40 | -1.21 | 0.31 | -1.84, -0.59 | -3.05 | 0.18 | -3.42, -2.68 | -0.016 | 0.003 | -0.022, -0.010 |
| Race/ethnicity | | | | | | | | | | | | | | | |
| Non-Hispanic Black | 0.09 | 0.41 | -0.75, 0.92 | -1.86 | 0.35 | -2.58, -1.14 | 0.68 | 0.53 | -0.39, 1.75 | 1.45 | 0.22 | 1.00, 1.89 | 0.027 | 0.006 | 0.014, 0.040 |
| Hispanic | -1.24 | 0.45 | -2.14, -0.34 | -0.14 | 0.32 | -0.78, 0.51 | -0.43 | 0.44 | -1.32, 0.47 | -0.42 | 0.27 | -0.98, 0.13 | 0.001 | 0.007 | -0.014, 0.016 |
| Non-Hispanic other race | 0.59 | 0.60 | -0.62, 1.80 | -0.53 | 0.62 | -1.78, 0.72 | 0.48 | 0.91 | -1.37, 2.32 | 0.89 | 0.34 | 0.20, 1.58 | 0.037 | 0.011 | 0.014, 0.059 |
| Educational attainment | | | | | | | | | | | | | | | |
| Less than high school | -0.13 | 0.41 | -0.96, 0.70 | 0.08 | 0.38 | -0.69, 0.86 | 1.16 | 0.58 | -0.01, 2.33 | 1.67 | 0.21 | 1.25, 2.09 | 0.017 | 0.005 | 0.006, 0.027 |
| High school diploma/GED | 0.04 | 0.34 | -0.64, 0.73 | 0.15 | 0.26 | -0.38, 0.67 | 0.67 | 0.43 | -0.20, 1.55 | 1.31 | 0.18 | 0.95, 1.68 | 0.012 | 0.004 | 0.003, 0.021 |
| Some college | -0.17 | 0.33 | -0.83, 0.49 | -0.15 | 0.26 | -0.69, 0.38 | 0.68 | 0.38 | -0.09, 1.44 | 1.15 | 0.22 | 0.71, 1.59 | 0.011 | 0.005 | 0.001, 0.020 |
| BMI category | | | | | | | | | | | | | | | |
| Overweight (≥ 25 to < 30) | -0.60 | 0.33 | -1.28, 0.07 | -0.13 | 0.32 | -0.77, 0.52 | -0.20 | 0.42 | -1.04, 0.64 | 0.05 | 0.21 | -0.38, 0.48 | 0.002 | 0.004 | -0.007, 0.011 |
| Obese I (≥ 30 to < 35) | 0.12 | 0.40 | -0.69, 0.92 | 0.28 | 0.35 | -0.43, 1.00 | 0.22 | 0.42 | -0.63, 1.06 | 0.55 | 0.23 | 0.08, 1.03 | 0.012 | 0.004 | 0.004, 0.020 |
| Obese II (≥ 35) | 0.71 | 0.51 | -0.31, 1.74 | 1.15 | 0.38 | 0.39, 1.91 | 1.86 | 0.52 | 0.81, 2.91 | 1.64 | 0.26 | 1.12, 2.16 | 0.033 | 0.005 | 0.022, 0.044 |
| Smoking status | | | | | | | | | | | | | | | |
| Current | 0.14 | 0.49 | -0.86, 1.14 | 0.68 | 0.32 | 0.03, 1.33 | 0.96 | 0.47 | 0.01, 1.91 | 7.37 | 0.28 | 6.79, 7.94 | 0.122 | 0.007 | 0.108, 0.136 |
| Former | 0.14 | 0.28 | -0.42, 0.71 | 0.00 | 0.23 | -0.47, 0.47 | 0.25 | 0.27 | -0.30, 0.81 | 1.97 | 0.14 | 1.68, 2.26 | 0.026 | 0.003 | 0.019, 0.032 |
| Alcohol use | | | | | | | | | | | | | | | |
| 1-4 drinks per day | -0.08 | 0.26 | -0.61, 0.44 | -0.41 | 0.22 | -0.86, 0.04 | -0.09 | 0.34 | -0.77, 0.59 | -0.25 | 0.14 | -0.54, 0.03 | -0.006 | 0.004 | -0.013, 0.001 |
| 5+ drinks per day | 1.52 | 0.80 | -0.10, 3.14 | 0.52 | 0.66 | -0.82, 1.87 | 2.58 | 0.79 | 0.98, 4.19 | 1.34 | 0.55 | 0.24, 2.44 | 0.003 | 0.011 | -0.019, 0.025 |
| *N* = 3,473 | | | | | | | | | | | | | | | |

Note. PoAm = Pace of Aging methylation; *CI* = confidence interval. Bold font denotes statistically significant association following false discovery rate correction.
